# Professional experiences on use of the mental health act in ethnically diverse populations: a photovoice study

**DOI:** 10.1101/2024.10.13.24315024

**Authors:** Kamaldeep Bhui, Roisin Mooney, Doreen Joseph, Rose McCabe, Karen Newbigging, Paul McCrone, Ragu Raghavan, Frank Keating, Nusrat Husain

## Abstract

**Background:** There are long standing ethnic and racial inequalities in experiences and outcomes of severe mental illness, including compulsory admission and treatment (CAT).

**Objectives:** To gather professional experiences about a) remedies for ethnic inequalities in the use of the Mental Health Act (1983, 2007;MHA); b) recommendations for improving care experiences and for reducing ethnic inequalities.

**Method:** We undertook a participatory research process using photovoice to gather experience data. Photographs were assembled and narrated by 17 professionals from a variety of disciplines. We undertook thematic analysis.

**Results:** Ineffective communications between inpatient and community services, insufficient staff capacity, a lack of continuity of care, and language and cultural constraints meant MHA assessments were lacking information leading to elevated perceptions of risk. Practitioners felt helpless at times of staff shortages and often felt CAT could have been prevented. They felt voiceless and powerless, and unable to challenge stereotypes and poor practice, especially if they were from a similar demographic (ethnicity) as a patient. Interdisciplinary disagreements and mistrust led to more risk aversive practices. The legislation created an inflexible, risk averse and defensive process in care. Police involvement added to concerns about criminalisation and stigma. There were more risk averse practices when team members and families disagreed on care plans. More rehabilitation and recovery orientated care is needed. Legislative compliance in a crisis, conflicted with supportive and recovery orientated care.

**Conclusion:** Clearer standards are needed including specific protocols for MHA assessment, police interactions, considering alternatives to admission, early intervention and continuity of care.

**What is already known on this topic:** The Levels of compulsory admission and treatment (CAT) are rising, and ethnic and racial disparities in level of detention persist. Practitioners’ experiences of using mental health legislation holds valuable information on how to reduce ethnic and racial disparities, yet their views are rarely sought for innovations in policies and practice.

**What this study adds:** Practitioners identified several reasons why ethnic and racial disparities persist, including increased risk perception due to interdisciplinary mistrust and poor communication, family discord, and a lack of information due to language and cultural constraints. Police involvement led to escalation, and police and mental health practitioner roles were not always clear, leading to frustration. The emotionally demanding work of caring for people with severe mental illness, and then undertaking CAT often involved disagreement and lead to fatigue and failures to consider all options at the time of assessment. Defensive practice, delays in assessment, a lack of continuity of care, and staff shortages all add to imperfect decision making and escalation to CAT rather than other options.

**How this study might affect research, practice or policy:** Improving legislation alone will not reduce ethnic and racial disparities in CAT, rather a comprehensive range of community services, skilled interdisciplinary communication and decision making, less escalation to police involvement, and tackling staff shortages are all essential. Culturally competent care also requires better skills in assessing across language and cultural barriers, as well as involving family in decisions.

## Background

In Europe and North America, ethnic minorities and migrants experience more adverse pathways to mental health care, including higher rates of compulsory admission and treatment (CAT), more contact with the police and criminal justice agencies, as well as poorer long-term outcomes compared with White British people. ^1–5^ ^6–8^ CAT increases stigma, fear, avoidance of mental health services, and feelings of powerlessness, and undermines autonomy, agency, and recovery among patients. ^9–12^ Patients find CAT distressing and frightening, especially due to the use of force and restraint and seclusion, and loss of autonomy; providing clear information in the context of collaborative relationships can be helpful.^13^ Black people in particular feel decisions are made by others, and they are mistreated, although they also acknowledge admission can be helpful.^14^

An Independent Review of the Mental Health Act (MHA England & Wales) made recommendations that might reduce ethnic inequalities in CAT. It also recommended more experience-based research especially from service users and professionals involved in applying the MHA. Research can expose mechanisms and processes that result in higher detention rates in some groups and not others, and inform how legislation and care might be improved, alongside actions to reduce the use of CAT. ^1^ ^2^ Lived experience expertise is beginning to challenge conventional paradigms of research and care through new insights, however, professional perspectives are also important as these too can provide important insights. Not least, professionals involved in implementing the MHA will have knowledge gathered over many years of practice, from different settings and for many different populations. This study reports on professionals’ experiences of MHA use in ethnically diverse populations.

## Aims

To assemble the experiences of professional stakeholders from multiple disciplines to:

a. Offer potential solutions to reduce ethnic inequalities in CAT..
b. Make recommendations to improve legislation, practice and policy, and support people subject to CAT.

## Methods

### Methodological Innovation

Understanding what drives inequalities is challenging. Qualitative research offers a powerful approach to discover novel interacting drivers of inequalities, for example, it has been used to explain clustering of diabetes, depression and HIV by ethnicity and gender.^15–17^ ^18^ Crenshaw’s intersectional approach ^3^ ^18^ ^19^ in particular exposes how inequalities arise and persist taking account of multiple co-existing influences. Gathering experiential data presents its own challenges, as people vary in their willingness and ability to recount events, especially if these are experienced as stressful, traumatic, or if they fear criticism and allegations of poor practice. We were guided by a systematic review of participatory methods, which identified photovoice (PV) as a promising approach for such situations.^20,1^ PV engages participants in a creative and reflective process of perspective taking, permitting iterative deepening and exploration of narratives. PV disrupts the traditional power relationships between researcher and participant, and can expose intersectional mechanisms.^21^ ^22^

### Setting and Participants

Participants were invited to three workshops: a first consisted of an induction session, providing information about the study and then sought consent to participate. In the second, participants shared photographs.. Reflections emerged in discussion prompted by the images, and participants developed short descriptors for their images (captions). In a third workshop, participants came together to review the data (including captions, summaries and images) to deepen their reflections and identify key moments where different decisions and actions might have changed or improved their experiences and patient outcomes.

Of seven potential recruitment venues (London, Birmingham, Manchester, Leeds, Bradford, Oxford, Derby, and Lancashire), participants were from Leeds, Derby, Oxford, and Lancashire. Pressures on clinical time and the time needed to undertake the activity were partly responsible for not recruiting from all sites. The professionals entering the study were from diverse ethnic groups: White British (N=11), Pakistani (N=3), Sri Lankan (N=1), African (N=1), and Indo-Caribbean (N=1). They included psychiatrists (N=3), nurses (N=4), psychologists and psychotherapists (N=3), occupational therapists (N=2), social work (N=1), and managers (N=4). There were ten men and 7 women, with ages ranging from 26-51; career durations in these roles ranged from 1 month to 15 years.

### Data analysis

The data consisted of photos, captions, transcripts, and field notes, managed in MAXQDA. The initial analysis involved coding personal experience, narratives, pictures, and captions from professionals in the workshops. We carried out a thematic analysis, as described by Braun and Clarke^23^ as a method for identifying, analysing and reporting patterns within data. We also drew on the principles of polytextual thematic analysis, where participants also considered the image data for meaning and context and the starting point for the sentiments that were narrated.^24^ We undertook the following steps: view images and captions, read transcripts of emergent narratives, familiarisation, coding, searching for common themes and important uncommon themes, reviewing and refining themes, reporting.

The study was granted REC and HRA approval [21/SC/0204], and local NHS research department approvals.

## Results

### Images and Captions

We present a selective number of images and captions to illustrate some of the common themes that emerged as well as uncommon reflections that seemed relevant for our objectives (see Figure 1). The images and captions allude to distress at seeing people detained, concern about children of parents being detained, the care of pets who are often left behind, the messy and prickly nature of relationships and communications, silencing of the person (patient and professional in various contexts), as well as how much *mess* was left following a MHA assessment and CAT. The demographics (ethnicity, age, location, role) of the people producing the pictures are colour coded (key in the last image).

**Figure 1:**
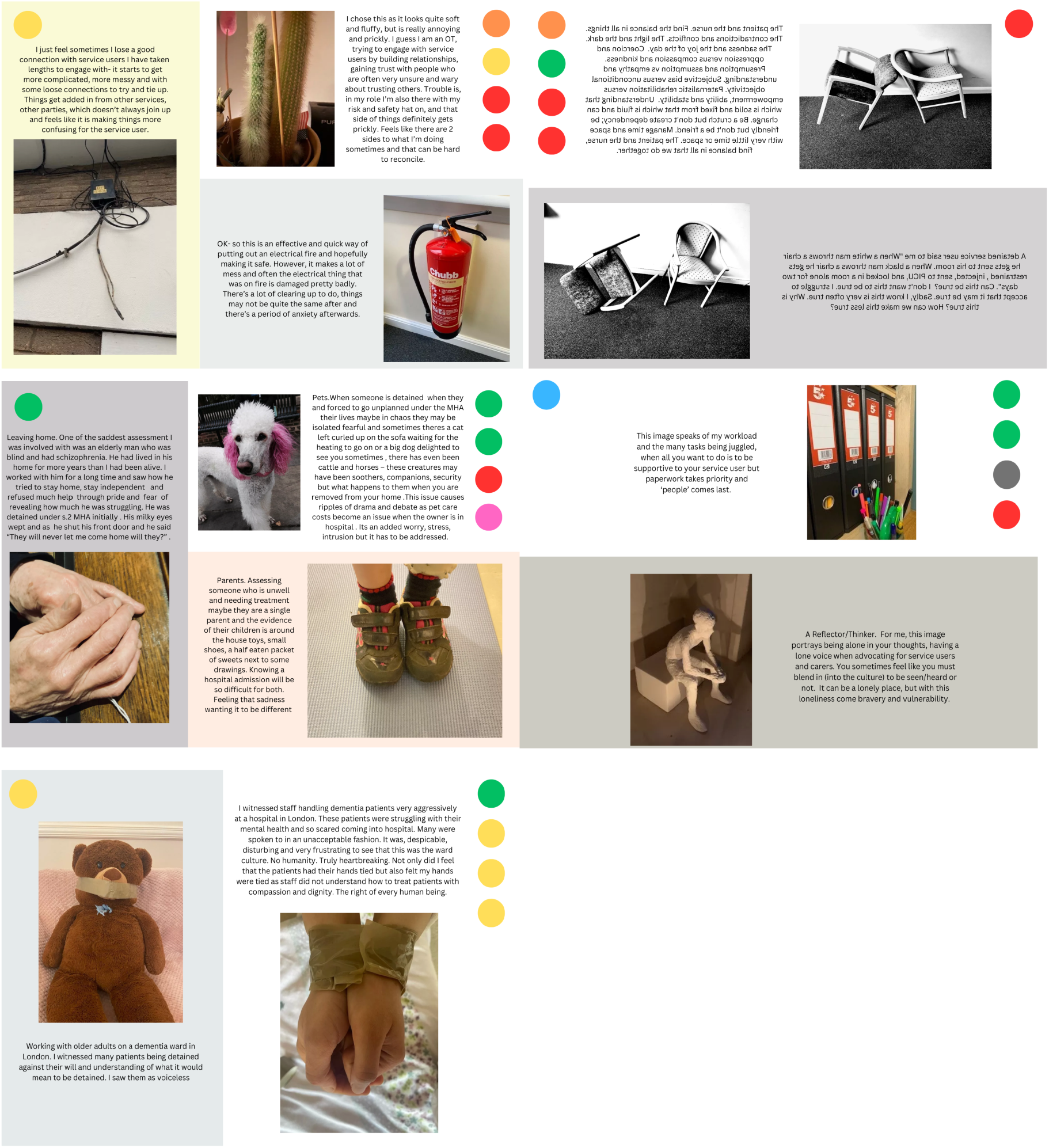
Professionals Photovoice Images and Captions

### Coding the dating and consistency

We identified 322 individual detailed codes of which 305 overlapped between two coders; these were verified by a third reviewer, and differences reconciled. Higher order themes were inductively generated, grounded in the data, in the synthesis. Below we present our findings, *italicising* elements that suggest *mechanistic* targets for improving care quality and prevention of CAT.

There were three high level themes (See Table 1 for some direct quotes and Table 2 for sub-themes):

1. Service operation and care pathways
2. Professional capabilities, skills, attitudes and experiences
3. Societal, health, and social structures

**Table 1:**
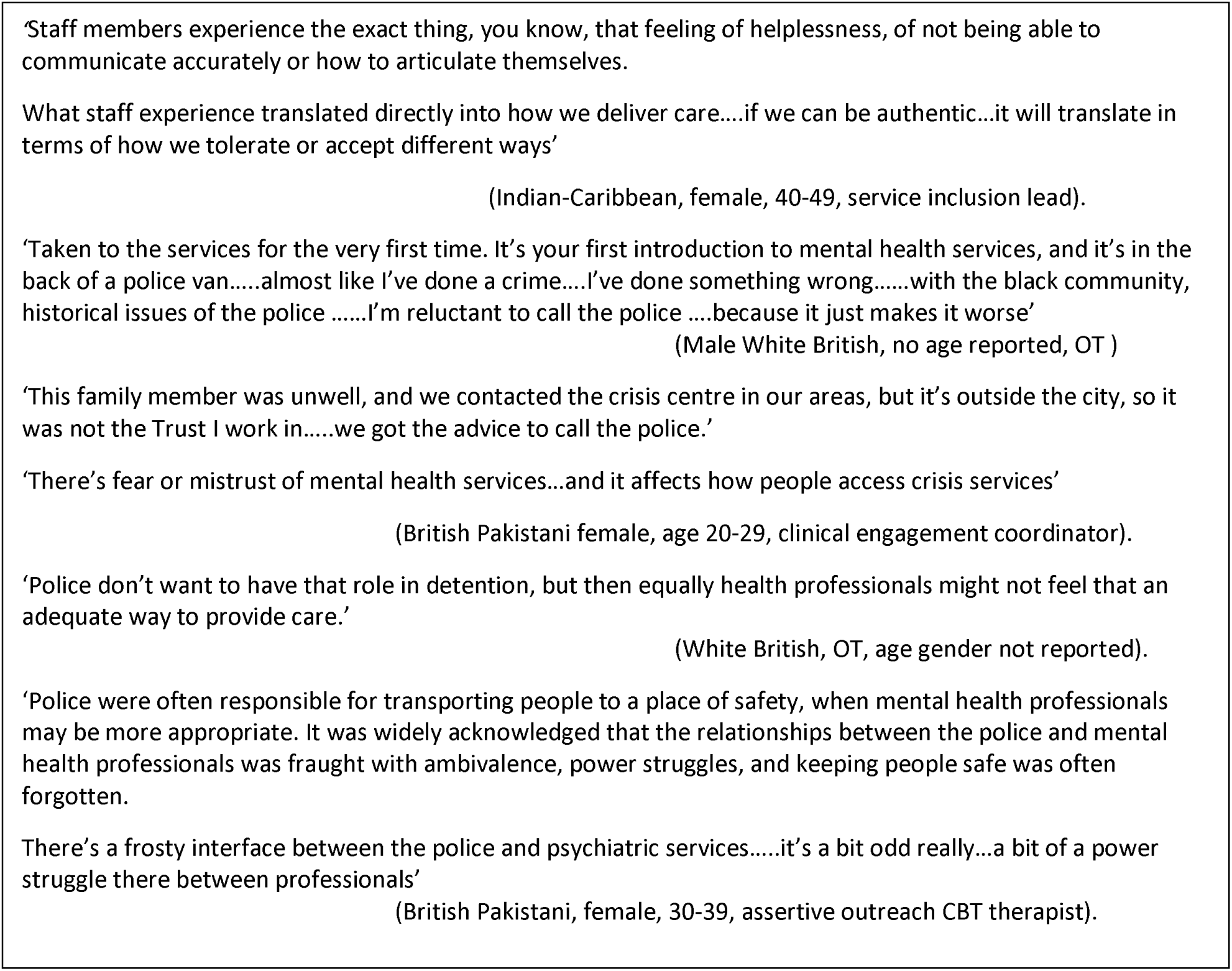
Specific Quotes.

**Table 2:**
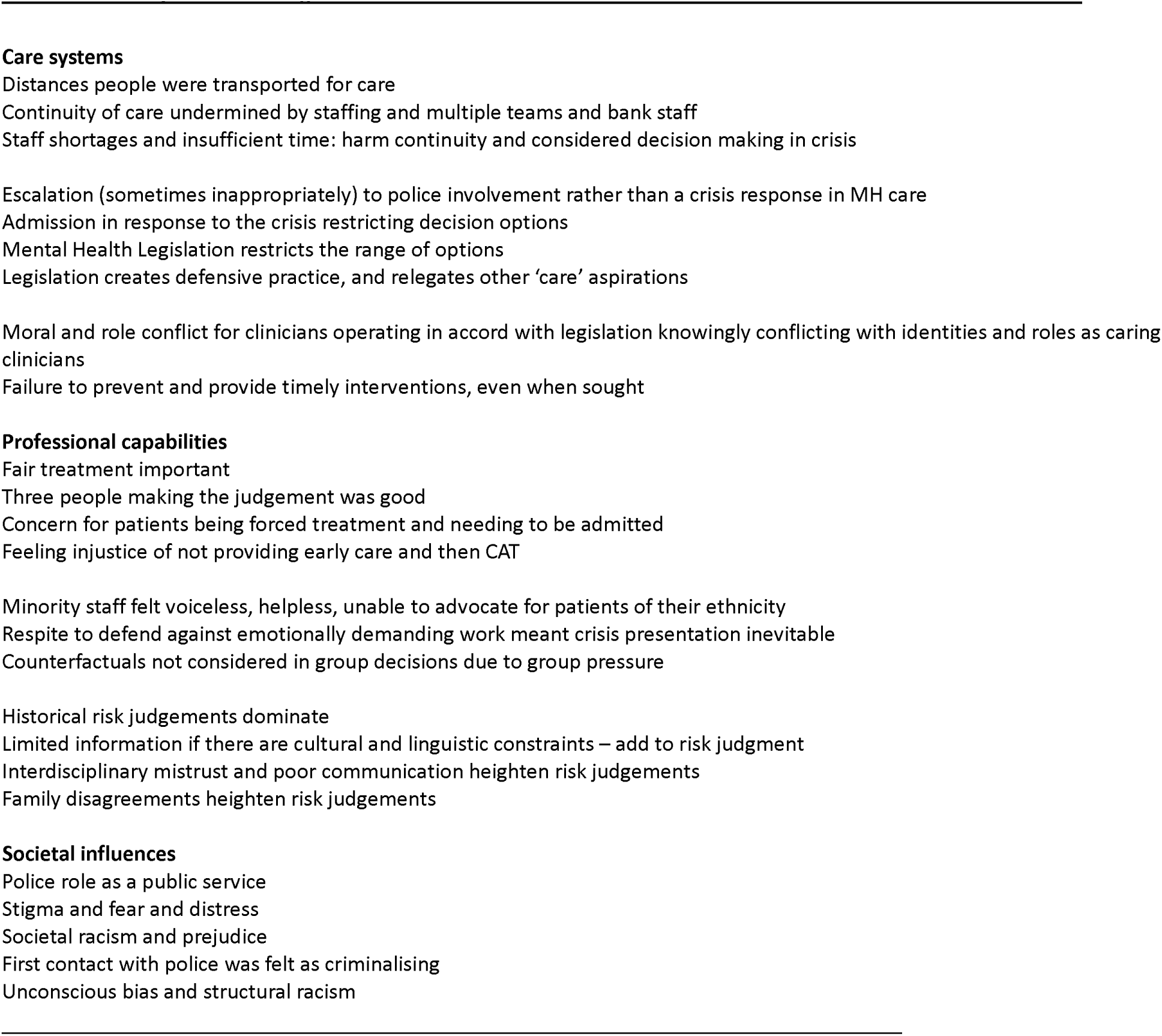
Summary of Critical Insights.

### Services operations and care pathways

There was consistency in narratives and themes across participants. Specifically, patients were sometimes *transported far from home*, and this was not helpful for recovery and made family visits difficult. *Large caseloads* prohibited more time being spent on individual patient care. *Limited time* meant *concerns were not worked through and resolved* during MHA assessments; crises thus ended up a self-fulfilling prophecy and CAT was recommended.

There was no *continuity of care* due to the number of teams and interfaces between different services, the very common use of ‘bank’ staff (non-regular staff working to cover vacancies and uncovered shifts), and many different staff interacting with patients even in one service. Assessments under the MHA were limited when there were *cultural and language constraints*, leading to *elevated perceptions of risk*; more nuanced contemporary information was not always available or gathered to better inform a more personalised and contextualised decision. *Risk assessment was then dominated by historical incidents*, rather than being based on more recent and contemporary information. Therefore, s*tereotypes* came into play. There was an acceptance of *unconscious bias* (e.g. if someone throws a chair it will be *differently interpreted done by a black* man rather than white man). A white middle-aged male participant explained how if a black person walked back and forth looking agitated, the tendency was to anticipate something ‘kicking off’.

Staff acknowledged that help for patients was often *offered too late* to avert a CAT episode. *People were refused help* when they sought care, and ended up in crisis, and then were forced to have care, as if it was inevitable or their fault. Participants were left feeling this was unjust. Participants felt that assessments *attempted to be fair*, in that there were *three people* making a judgement about suitability for detention, and this was a built-in opportunity to question. However, *counterfactuals were rarely entertained*, and it was uncommon to consider all options, suggesting *group pressures* lead to decisions that were not critically considered. Discontinuous care, insufficient time to work with patients in the community, or consider all the information, all added to perceptions of elevated risk, along with a failure to act early, and so each added to likelihood of a CAT episode following assessment.

### Professional capabilities, skills, attitudes, experiences

Professionals from the same ethnic or racial demographic as the patient felt *voiceless, isolated*, unable to speak up on behalf of the patient, and felt *helpless* themselves to change the prevailing views and ways in which decisions were weighed.

There were *conflicts in expected and experienced roles*, for example, between a person who wanted to provide good care versus *care being limited to following the MHA procedures.* Participants felt assessments then *focused on risks*, *anticipating blame,* and *fears about adverse outcomes*, such as a fatality. CATs happened even it was felt to be unhelpful because of defensive practice and role strain. Professionals felt unable to take the risk of not admitting in crisis, one person citing an example of a death following non-admission. Participants were saddened when people asked for help and were turned away, and then they ended up being forced to receive crisis treatment, which was distressing through no fault of their own.

Participants acknowledged the *importance of treating people well and fairly* and did not feel they always achieved this. Interestingly, multidisciplinary decision making was proposed to be important, to improve risk assessment and care planning. However, a lack of *interdisciplinary skills and mutual trust* made people risk averse and a CAT episode was more likely in such instances. Furthermore, there was *insufficient risk dialogue with patients*, who did not understand how judgements about them were being constructed, and why they were detained and might be secluded.

The work was *emotionally demanding*, and especially if there were disagreement with families, or when staff themselves felt under pressure (time, risk, powerlessness). They described a process of passively accepting *disengagement for respite* knowingly awaiting the next crisis. This was described a form of *resting away from the pressures of work* and demands placed upon them. People were very aware they might be *re-enacting a rejection* from an early phase of life for the patient, through the application of standard procedures. In this context, when there were *communication breakdowns* with patients, it was not easy to restore connections. There were interesting recommendations to try and *connect through sharing films, or books, or through music*, and *learn something about the person beyond their illness*.

### Societal, health, and social structures

The participants acknowledged wider societal issues, and how mental health care was represented in the public imagination: stereotypes fed perceptions, and stigma; they argued that we need better recovery orientated dialogue, and commensurate changes in society. Prejudice was proposed to be built into all processes, racism inevitably played a part, and therefore patients did not seek help.

## Discussion

### The Context

Mental health legislation permits CAT in many countries around the world; the legislation is intended to codify when and how patients may be detained. In so doing, its purpose is to protect the human rights of the patient, whilst guiding clinicians to follow transparent and legally specified processes that include safeguards. However, there is much dissatisfaction with mental health legislation as recommended procedures are not always followed.^25^ Ethnic inequalities in the use of CAT raise concerns about bias in assessments and judgements about risk. They raise concerns about societal and institutional racism and whether the wider social determinants and systems of care are racially, ethnically, and culturally unfair. Risk assessment is rarely predictive and is bedevilled by lack of agreement between assessors, and unconscious bias.^26–28^ ^29^ Furthermore, relatives views may not align with those of patient or professionals creating uncertainty at a time of crisis.^28^ The Independent Review of The Mental Health Act was firm that legislation alone was not going to reduce ethnic disparities in CAT, and investment in a comprehensive care system was essential for any legislation to be useful and helpful.^30^ ^31^ Professionals who work in the care system are likely to be aware about what explains these inequalities.

### The Findings

This study sought to gather professional perspectives on how to prevent CAT and improve care experiences. Our findings show significant points of intervention (see Table 2). These can be addressed by improved professional standards and training and care systems reforms, and some may be mitigated by advanced agreements and advocacy. A recent evidence synthesis sought to explain rising detentions in the UK, proposing a new explanatory model for rising CAT.^32^ Our findings confirm some of the proposed mechanisms reported in that evidence synthesis:

- Defensive practice and concerns about the consequences of not admitting in crisis
- Crisis and MHA assessment occurred too late, and there was little critical thinking, or evaluation of alternatives and consideration of counterfactuals
- Lack of continuity of care and insufficient community support
- Staff shortages, the number of teams, bank or non-regular staff, all add to the lack of continuity

Our findings add additional mechanisms:

- Interdisciplinary mistrust and lack of effective communication increases perceptions of risk
- Family discord increases perceptions of risk
- Police involvement led to escalation rather than prevention, ultimately ending in CAT
- Unclear roles and responsibilities between the police and mental health professionals make for imperfect decisions and frustrated relationships.
- Emotionally demanding work of MHA legislation and care for people when there is disagreement can lead to fatigue and failures to consider all options
- Lack of sufficient information due to language or cultural constraints leads to perceptions of greater risk
- Professionals (especially those from minority ethnic groups but generally also) are unable to object to group pressures, and feel isolated and without voice
- MHA assessments prioritise the legal process and compliance, forcing codified decision options rather than a fuller range of personalised options

Research of interventions and quality improvement strategies targeting these mechanisms may be able to prevent admission by strengthening actions earlier in the care pathway, as well as encouraging *critical and resistant* thinking at the time of the assessment. This may require longer assessment, consultation with interpreters and family, and GP. There is also a need for skills in interdisciplinary working and ensuring mistrust and poor communication do not undermine optimal and least restrictive options.

### Structural Analysis

The findings draw attention to wider societal views, including stigma, prejudice, historical relations of police and ethnic minorities, all driving fear of services and undermining trust. These are important for two reasons; first the lack of trust may not be overtly expressed but may lead to fear and conflict, and a need for greater attention to how to address mistrust in a crisis. The second is that professionals identified some people who would not benefit from admission, yet they felt compelled to follow legislative procedure, somewhat dispassionately; in response they felt a moral and ethical conflict in doing so. Professionals too were distressed. The professionals echo what patient have said for some time, and both seem entangled in a system that works for neither.^33^ ^13^ How does this happen?

Bourdieu introduced an important concept of ‘habitus’, which Lo and Stacey elaborated to understand cultural interactions in clinical care.^34^ Habitus links cultures, contexts, and social structures, thus the clinical encounter is a product of the health/social system, operating within the social structures and broader values that condition what takes place at all levels. Therefore, traditions are reproduced over time to functionally adapt but these then sustain the very challenges and harms that progressive policy and practice seek to mitigate. Thus, there is a need for resistance and critical reflection, which is hard in a crisis.

We previously used the notion of habitus in understanding how to improve cultural competency through a cultural consultation model of training in East London, comprising clinical ethnography and assessments of identity, explanatory models, psychosocial factors, and therapeutics.^35^ Creating less crisis orientated spaces in which to consider relevant factors and appreciate the power of institutional forces may offer more opportunities for critical thinking, and resistance to group, institutional, and legal pressures.^35^ ^36^

MHA assessments may be psychologically traumatic for the carers and practitioners who may show signs of hypervigilance, desensitization, disillusionment, and distrust that might add to poorer communication, exhaustion, fear of adverse consequences, and disempowerment.^37^ All of these can increase defensive practice and risk perceptions, ironically, at a time when a patient is likely to be terrified and feel entirely powerless. Escalation to involve the police rather than apply a preventive intervention already signals a lack of therapeutic connections and speaks to the historical and ongoing power of institutions to criminalise.

The interplay of power and poverty, and long-standing epistemic injustice continues to face people with mental illness due to the stigma and negation of their views on grounds that they do not have insight. Professionals too expressed a sense of not being heard nor being able to speak up.

Some argue mental health legislation and coercive care are incompatible with human rights and the interest of those living with disabilities.^38^ Previous legislative reviews claimed the MHA (England and Wales) appears to violate UN Convention on the Rights of Persons with Disabilities (CRPD), for example, Article 4 (‘no discrimination of any kind on the basis of disability’), Article 12 (persons shall ‘enjoy legal capacity on an equal basis with others in all aspects of life’), and Article 14 (‘the existence of a disability shall in no case justify a deprivation of liberty’).^39^ However, the realities of daily practice are that legislation is required in instances to protect against harms to the individual or others, but its application requires more precise and contextualised judgements, and a broader range of options and alternatives to admission.

## Conclusions

We evidence new mechanisms by which ethnic inequalities of CAT take place and how this might be prevented. We propose that health systems must ensuring sufficient capacity and continuity of care, the required range of specialist services so early and preventive intervention is the norm, and alternatives to CAT. New legislation is likely to be optimally effective only if these conditions are met. Advanced agreements and advocacy seem especially relevant and potentially helpful as an opportunity to prevent or critically plan for preferred care options in future crises. A notable finding is that police involvement was not always necessary but did add to fear and stigma, and clarity is needed on their future role given the history of racism and police interactions and concerns about how mental health services are perceived if there is a reliance on the police. Professional perspectives and demoralisation signal a failure of the system and conflict with professional values. Unless there is widespread service and systems reform, it is likely MHA will continue to be used as a default when other parts of the system do not operate well. A legislative option is not ideal for the patient or the professional.

## Conflicts of interest

KB is an associated editor at BMJ Mental Health but played no part in the processing and decision making on this paper.

None otherwise declared.

## Contributions

KB and RM were PI and co-PI on this NIHR funded study. KB drafted the paper, based on data collected by RM and other members of the research team. KB analysed the data, and all authors contributed to interpretation and revision of consecutive drafts.

## Ethical Approval

The study was granted ethical approval from the National Health Service (NHS) Research Ethics Committee (REC) and Health Research Authority (HRA) on the 1st December 2021, after review at the South Berkshire Committee [21/SC/0204].

## Data Availability

This is qualitative data set, a small sample, with unique identifiers and descriptors as is usual in qualitative research; we have presented only intersection characteristics; therefore the data set is not easily re-analysed by other parties. We have not made the data available given the risk of identification by locality and other professional and demographic characteristics.

